# Livestock production intensity and mucosal IgA and IgG responses to H5N1 highly pathogenic avian influenza A virus, North Carolina, 2021-2022

**DOI:** 10.64898/2026.08.06.26359901

**Authors:** Nora Pisanic, Kathleen M. Kurowski, Timothy Carter, Bonita D. Salmerón, Kristoffer Spicer, Kate L. Kruczynski, Carolyn Gigot, Matthew A. Aubourg, Laura Schmidt, Devon J. Hall, Devon J. Hall, Leroy Mitchell, Linda Johnson, Myranda George, Ana M. Rule, Meghan F. Davis, Gigi K. Gronvall, Andrew Pekosz, William J. Moss, Christopher D. Heaney

## Abstract

**Background:** Direct livestock exposure is a risk factor for zoonotic influenza, including H5N1 highly pathogenic avian influenza (HPAI) A virus. But whether living in regions of high poultry and swine production intensity (PPI, SPI) increases risk of exposure to zoonotic influenza viruses independent of occupational livestock contact remains unclear.

**Objectives:** To determine whether livestock workers and community members with no occupational livestock exposure in North Carolina, where poultry and swine production are increasingly co-located, are at higher risk of exposure to zoonotic influenza.

**Methods:** Saliva samples from industrial livestock operation worker (ILO-W), ILO neighbor (ILO-N) and metropolitan area (Metro) households were analyzed for mucosal influenza A (H5N1, H1N1, and H3N2) hemagglutinin (HA) IgA and IgG antibodies to determine associations of PPI, SPI, exposure group, and detection of a swine-specific fecal contamination marker (Pig-2-Bac DNA) with influenza A antibody levels.

**Results:** Residing in the highest PPI and SPI tertile was associated with significantly higher mucosal H5 and H1 HA IgA levels, including among residents without occupational livestock exposure. Households with occupational poultry or swine contact had significantly higher H5 IgA and IgG and H1 IgA levels compared to Metro households. In regression models accounting for clustering at the participant level, log_10_ anti-H5 HA mucosal IgA increased 0.16 (95% CI: 0.06, 0.27, *p*<0.005) and 0.10 (95% CI: 0.03, 0.17, *p*<0.005), per log_10_ increase in PPI and SPI, respectively, and 0.16 (95% CI: 0.03, 0.19, *p*<0.02) when Pig-2-Bac DNA was detected on household surfaces.

**Conclusions:** Mucosal H5 HA IgA and IgG and H1 HA IgA were consistently elevated across different metrics of livestock exposure intensity, including residential exposure, occupational contact within a household, and a molecular marker of household swine fecal contamination in a state with intensive poultry and swine production.

## Introduction

Highly pathogenic avian influenza (HPAI) A (H5N1) viruses have spread extensively among wild birds, poultry, and mammalian species in North America since 2021, increasing concerns about zoonotic transmission and viral adaptation to sustain human-to-human transmission [1–3]. Over 70 cases of human H5N1 infections have been documented in the United States, mostly associated with occupational exposure to infected poultry flocks or dairy herds, but the full extent of infection remains uncertain, as mildly symptomatic and asymptomatic patients may be missed by clinical surveillance [4–6].

During the 2024 U.S. H5N1 outbreak, H5 influenza RNA was detected in municipal wastewater systems across nine states. Among the sites that tested positive for influenza A H5, 37% (9/24 sites) could not identify any livestock, dairy or avian inputs within the sewershed [7]. The detection of H5 in municipal sewersheds without known input from livestock sources raises the possibility that occult infections of people in communities could be occurring. Other possibilities include environmental contamination of the sewershed or household consumption and disposal of contaminated raw milk or raw milk products.

Most zoonotic influenza risk research has focused on occupational exposure among poultry and swine workers and more recently dairy cattle workers [8]. Swine production systems are of particular concern because pigs are susceptible to both avian and mammalian influenza A viruses and may facilitate viral reassortment and emergence of novel strains with pandemic potential [9]. Intensive poultry production could also increase opportunities for environmental dissemination of avian influenza viruses through transport and spreading of poultry waste, contaminated dust, and aerosols [10]. In communities of rural eastern North Carolina, where poultry and swine production are increasingly co-located, the zoonotic influenza risk may be particularly high. But whether living in regions of high livestock production intensity increases risk of exposure to zoonotic influenza viruses independent of occupational livestock contact remains unclear.

Serologic approaches can identify prior influenza exposures that may not have been clinically recognized. Saliva-based antibody testing offers several advantages for community-based epidemiologic studies. Specimen collection is noninvasive and well suited for repeated longitudinal sampling. Importantly, mucosal IgA is increasingly recognized to play an important role in protection and fast recovery from respiratory virus infections, including influenza A [11] and SARS-CoV-2 infection [12]. Recent work has demonstrated the utility of bead-based multiplex immunoassays for measuring pathogen-specific antibody responses in saliva with performance characteristics comparable to blood-based serologic assays [13,14]. Innovations in multiplex immunoassay technology, including a dual reporter format, permit simultaneous assessment of antibody levels to multiple influenza antigens and isotypes (mucosal IgA and blood transudate IgG).

In this study, we evaluated mucosal influenza A virus IgA and IgG responses, including responses to HPAI H5N1 HA, among industrial livestock operation (ILO) workers, their household members, and among rural and urban community residents of North Carolina who reside at varying distances from areas of intensive livestock production. Antibody responses were measured using multiplex immunoassays optimized for saliva and built on our previously described bead-based integrated serology platforms [12,13,15]. The goal of this study was to determine associations of regional livestock production intensity and household contamination with swine fecal material, measured by microbial source tracking (MST) marker Pig-2-Bac DNA contamination, with antibody responses to H5N1 and other influenza A viruses among both livestock workers and non-occupationally exposed residents.

## Methods

### Study Design

This study leveraged self-collected saliva samples from a previously described study conducted from February 2021 to December 2022 in North Carolina among industrial livestock operation neighbor households (ILO-N group; n = 95), metropolitan urban households (Metro; n = 108) and among ILO worker households (ILO-W; n = 95) [14,16]. ILO worker households provided weekly saliva samples for up to 4 months (up to 17 saliva samples total). Household addresses were geocoded and poultry production intensity (PPI) and swine production intensity (SPI) values were assigned to each home using a summed inverse distance weighted approach, as described previously [17]. Briefly, the intensity was calculated based on permitting records using the distance between the household and operations as well as a metric for size of the operation – steady state live weight for swine operations and live bird count for poultry operations. At each study visit (n = 1 visits for ILO-N and Metro groups, n=17 visits for ILO-W group) participants self-collected two types of saliva samples, a gingival crevicular fluid swab (Oracol S14, Malvern Medical Developments, UK) and a passive drool sample [14]. As published previously, participants also collected up to four household surface settled dust samples (two indoor and two outdoor samples), to test via quantitative PCR (qPCR) for Pig-2-Bac DNA, a microbial source tracking (MST) marker of swine fecal contamination [17]. The Johns Hopkins Bloomberg School of Public Health (BSPH) Institutional Review Board (IRB) approved this study (IRB00014420).

### Multiplex Immunoassay

Gingival crevicular fluid swab samples, which are enriched with serum-derived IgG antibodies [18], were used to measure the IgG response to influenza virus antigens, whereas passive drool was used to measure mucosal IgA. An in-house multiplex immunoassay based on Luminex xMAP technology was used to determine antibody responses against HA from H5N1 (A/chicken/Ghana/AVL-763_21VIR7050-39/2021) (Sino Biological, Chesterbrook, PA), H1N1 (A/Michigan/45/2015)(H1N1)-pdm09-like (eEnzyme, Gaithersburg, MD), and H3N2 (A/Singapore/INFIMH-16-0019/2016/H3N2)-like virus (eEnzyme). Antigens were coupled to beads (5 μg antigen/million beads) and successful coupling was determined using sheep or human influenza antibody control sera (NIBSC, UK) or polyclonal HA-specific rabbit antibodies (eEnzyme), followed by detection using Phycoerythrin (PE)-labeled anti-species antibodies. The assay procedure followed previously developed protocols with minor modifications [19]. Saliva was centrifuged for 5 min at 20,000 g. All following reactions were performed in black 96-well plates with 50-μL reaction volumes. Samples were incubated for one hour on a plate shaker at 800 rpm at a 1:10 dilution in assay buffer containing 1,000 beads per antigen or control (beads conjugated with anti-human IgG, anti-human IgA and BSA). Each plate contained blank wells (assay buffer) for background fluorescence subtraction. PE-labeled anti-human IgG (1:250 in assay buffer) was used to detect influenza-specific IgG in saliva; biotinylated anti-human IgA (1:2,000, both Jackson Immuno Research, West Grove, Pennsylvania), followed by brilliant violet-labeled streptavidin (1:500, BD Biosciences) was used to detect mucosal influenza virus-specific IgA. Assay plates were read on a Luminex iFlex dual reporter instrument (Luminex, Autin, TX).

### Statistical Analysis

Blank subtracted net median fluorescence intensity (net MFI) was used for analyses. For the ILO-W group, which had provided samples for IgG analysis at multiple study visits, the mean MFI response across visits was calculated for each participant to obtain a cross-sectional dataset (one IgG/IgA measurement/participant), which was used to plot crude differences (figure plots). Influenza IgG and IgA responses by PPI and SPI tertile were visualized in R Studio (R version 4.5.2) using packages ggplot, and ggpubr. Statistical differences between PPI and SPI level and between exposure groups were determined using ggstatsplot with non-parametric Wilcoxon test to derive *p*-values [20] and verified using generalized linear models (GLM) with generalized estimating equations (GEE) with an exchangeable working correlation to account for clustering at the participant level (IgG only). Unadjusted univariate GEE GLM model fits were compared to GEE GLM models adjusted for age, sex, and household size, prior influenza vaccination, season and study month and combinations thereof. Model correlation structures and fits were compared and selected based on Quasi Information Criterion (QIC) and QICu values (see **Table S1 & Table S2**).

## Results

### Participant characteristics

A total of 258 households (n = 298 participants) participated in this study. Metro households constituted the largest study group (n = 95 households, 37%), followed by ILO-Worker (ILO-W) households (n = 84, 33%) and ILO-Neighbor (ILO-N) households (n = 79, 31%). As described previously, poultry and swine production intensity (PPI, SPI) values were assigned to each household based on the permitted steady state weight of each livestock type raised in industrial livestock operations within residential vicinity. PPI and SPI were highest in the ILO-N and lowest in the Metro group; see **Table 1** for additional household and participant characteristics.

**Table 1.** Household and participant characteristics stratified by study group, North Carolina, USA, 2021-2022.

| Study Population Characteristic | ILO-Worker | ILO-Neighbor | Metro | All Groups |
| --- | --- | --- | --- | --- |
| Participants, n (%) | 95/298 (32%) | 95/298 (32%) | 108/298 (36%) | 298/298 (100%) |
| Children, n (%) | 3/13 (23%) | 4/13 (31%) | 6/13 (46%) | 13/13 (100%) |
| Male, n (%) | 47 (49%) | 42 (45%) | 32 (30%) | 121 (41%) |
| Age in years, mean (SD) | 42 (13) | 50 (17) | 40 (16) | 43 (16) |
| Race / Ethnicity, n (%) |  |  |  |  |
| Black / African American | 84/95 (88%) | 82/93 (88%) | 90/106 (85%) | 256/293 (87%) |
| Hispanic/Latino/a/Latinx | 8/95 (8%) | 10/93 (11%) | 5/106 (5%) | 23/293 (8%) |
| White | 1/95 (1%) | 1/93 (1%) | 8/106 (8%) | 10/293 (3%) |
| Asian American | 0/95 (0%) | 0/93 (0%) | 2/106 (2%) | 2/293 (1%) |
| Other | 2/95 (2%) | 0/93 (0%) | 0/106 (0%) | 2/293 (1%) |
| Household size, mean (SD) | 2.7 (1.4) | 2.0 (1.3) | 2.2 (1.2) | 2.3 (1.3) |
| Sampling date, mean (range) | Jan. 2022 (April 2021-August 2022) | August 2021 (March 2021-August 2022) | October 2021 (March 2021-August 2022) | November 2021 (March 2021-August 2022) |
| Oral fluid samples, IgA, n (%) | 88 (33%) | 81 (31%) | 95 (36%) | 264 (100%) |
| Oral fluid samples, IgG<sup>†</sup>, n (%) | 87 (36%) | 74 (30%) | 82 (24%) | 243 (100%) |
| Oral fluid samples, IgG<sup>†</sup>, n (%) for GLM GEE models, n (%) | 402 (72%) | 74 (13%) | 82 (15%) | 558 (100%) |
| Current flu shot, n (%) | 35/93 (38%) | 25/95 (26%) | 35/106 (33%) | 95/294 (32%) |
| Swine CAFOs within 3 km of household, mean n (SD) | 2.97 (3.34) | 3.69 (5.18) | 0.07 (0.33) | 2.34 (3.89) |
| Poultry production intensity, log<sub>10</sub> birds/km<sup>2</sup>, mean (SD) | 2.46 (0.51) | 2.59 (0.29) | 1.71 (0.21) | 2.19 (0.53) |
| Swine production intensity, log<sub>10</sub> pounds/km<sup>2</sup>, mean (SD) | 6.25 (0.38) | 6.36 (0.29) | 5.01 (0.45) | 5.77 (0.75) |

*Note: ILO: Industrial livestock operation, SD: Standard Deviation. <sup>†</sup>Mean influenza A IgG responses by participant were used for crude figure plots; unadjusted and adjusted models with generalized estimating equations (GEE) were used for statistical analysis to account for clustering at the participant or household level.*

### Influenza A antibody responses by poultry and swine production intensity

Influenza H5 HA and influenza H1 HA IgA responses among adult community members living in the upper 33% (3^rd^ tertile) of livestock production intensity were significantly elevated compared to those living in the lowest 33% (1^st^ tertile) of livestock production intensity (H5 HA: *p*<0.001; H1 HA: *p*<0.01; **Figure 1**). This trend remained consistent when children were included in the analysis. Participants residing in the 3^rd^ tertile of PPI and SPI also had significantly higher anti-H5 HA IgA levels than those in the second tertile (*p*<0.05) (see Supplementary **Figures S1 & S2**). Importantly, even after excluding participants with occupational livestock exposure, H5 HA IgA levels remained significantly higher among those living in the highest PPI (*p*<0.01 between 2^nd^ and 3^rd^ tertile) and SPI tertiles (*p*<0.05 between 1^st^ and 3^rd^ and *p*<0.01 between 2^nd^ and 3^rd^ tertile) i.e., among community members who have no direct livestock contact, but live in areas of high livestock production intensity (**Figure S3 and Tables S3 & S4**).

**Figure 1.**
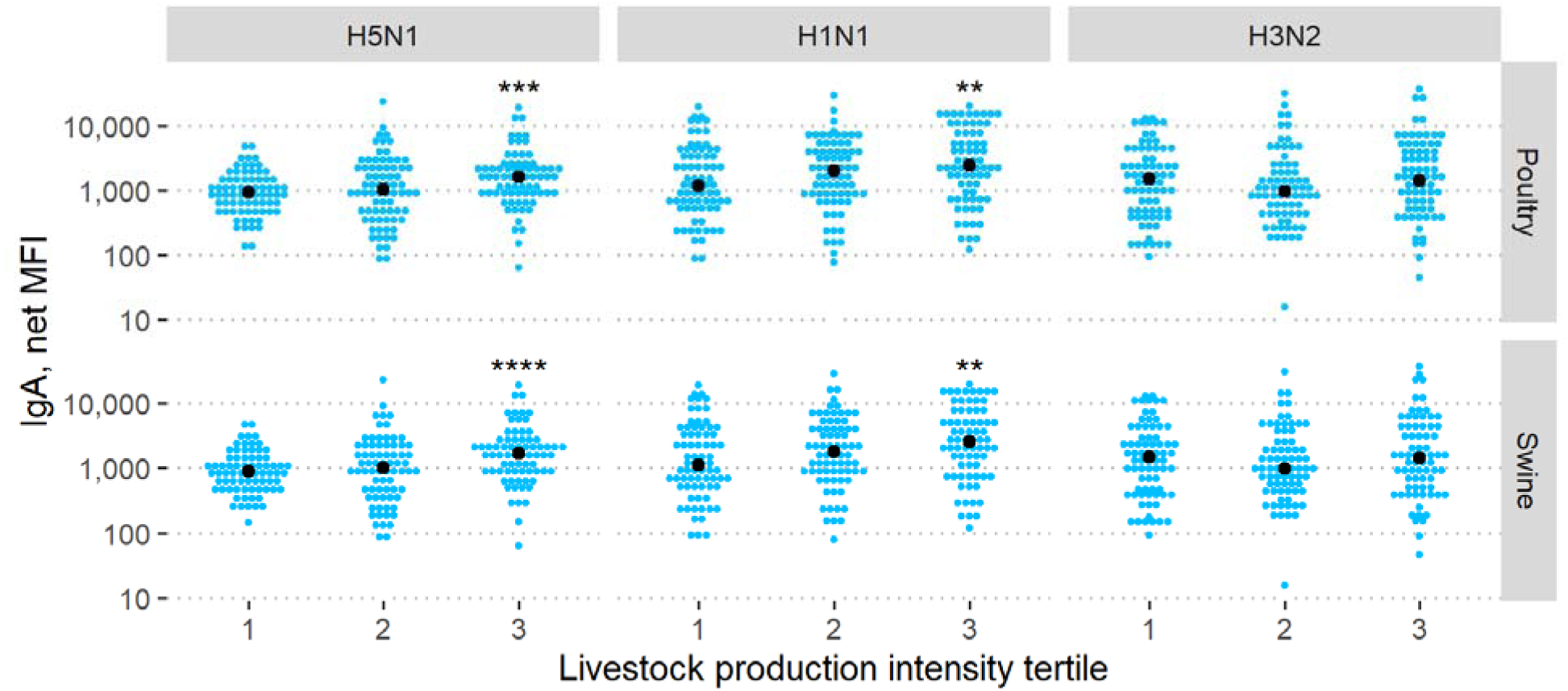
IgA responses to influenza A H1 HA, H3 HA, and H5 HA by livestock production intensity tertile (n=84 per tertile). Children (<18 years) were excluded from this analysis. Note: ** = p<0.01, *** = p<0.001, ****=<0.0001. Reference group for statistical comparison: 1^st^ tertile. MFI: Median Fluorescence Intensity

Similarly, IgG antibodies against H5 HA were also significantly elevated among community members living in areas within the highest livestock production intensity tertiles for both poultry and swine production (**Figure 2**).

**Figure 2.**
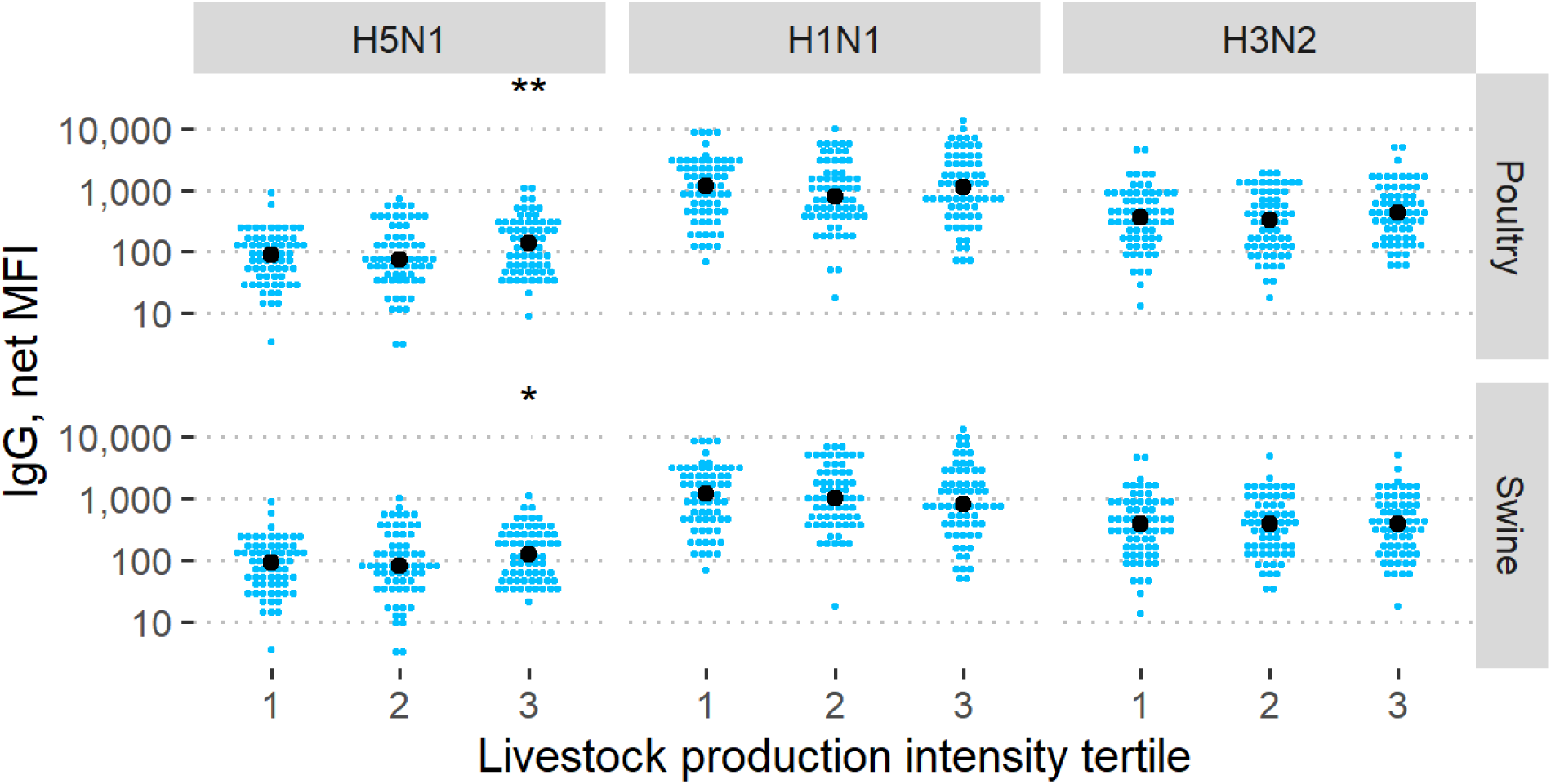
IgG responses to influenza A H1 HA, H3 HA, and H5 HA by livestock production intensity tertile. Children (<18 years) were excluded from this analysis. Note: *=<0.05, ** = p<0.01. Reference group for statistical comparison: 1^st^ tertile. MFI: Median Fluorescence Intensity.

### Influenza A antibody responses by household exposure group

When stratifying by study group, ILO-N and ILO-W households had significantly higher IgA levels to H5 HA and H1 HA than Metro households; ILO-W households also had significantly higher H5 HA IgG levels compared to Metro households with residences in urban areas of eastern NC with lower poultry and swine production intensity (**Figure 3**).

**Figure 3.**
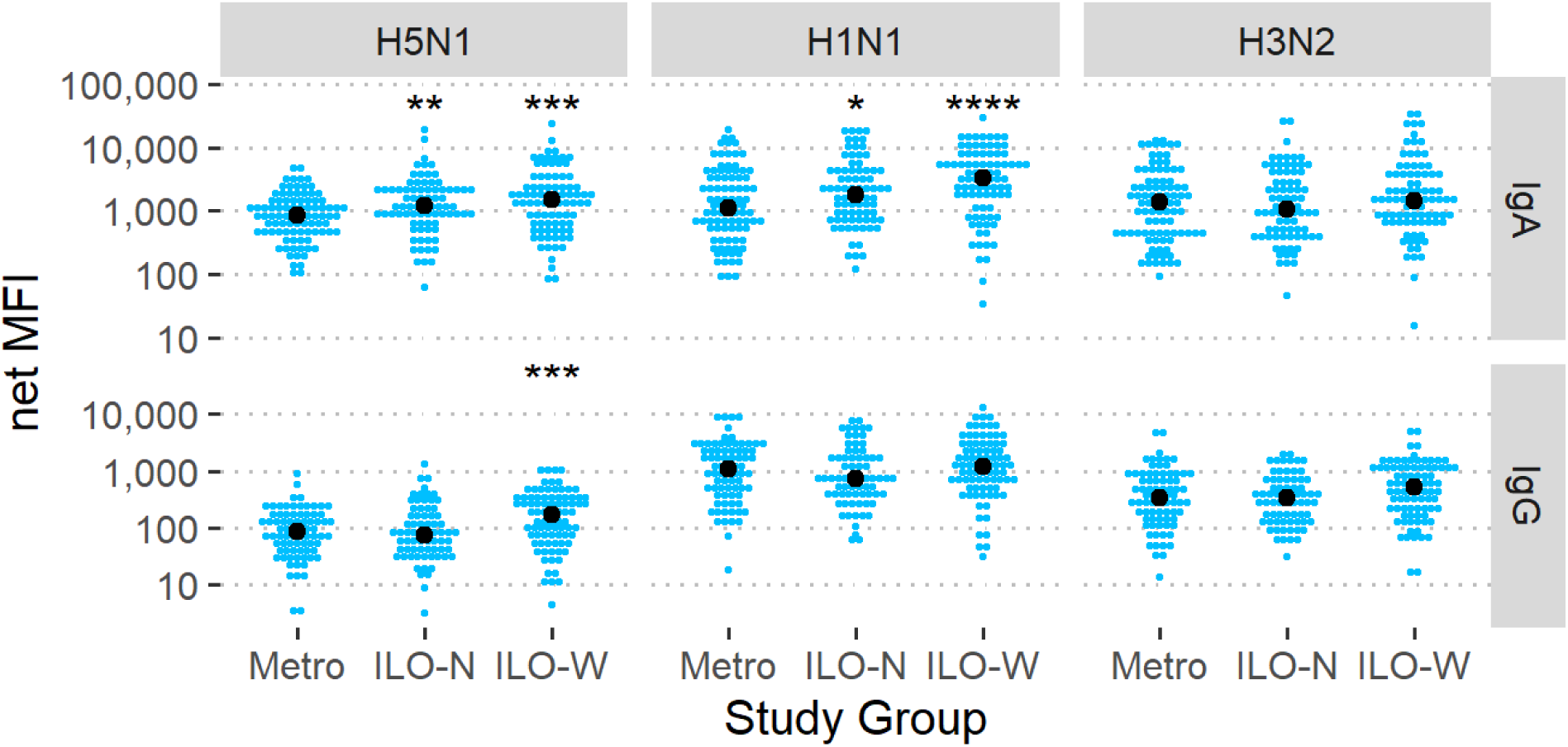
IgA and IgG levels specific to influenza A H1 HA, H3 HA and H5 HA stratified by study group. Note: *=p<0.05, ** = p<0.01, *** = p<0.001, ****=p<0.0001. Reference group for statistical comparison: Metro. ILO-N: Industrial Livestock Operation Neighbor Households, ILO-W: Industrial Livestock Operation Worker Households. MFI: Median Fluorescence Intensity

Comparing ILO-W households by occupational livestock species contact (predominantly poultry or predominantly hogs/pork) revealed that poultry exposure was associated with the highest H1 HA and H5 HA IgA and H5 HA IgG levels. Of note, households of industrial poultry operation workers also had significantly higher H3 HA IgA and IgG levels compared to all other groups (see **Figure S5** for *p-*values between additional exposure groups).

### Influenza A responses by household exposure to swine fecal material

We examined the association between detection of Pig-2-Bac DNA, a swine-specific microbial source tracking (MST) marker, and influenza A virus antibody responses. Residents of households where Pig-2-Bac DNA was detected on at least one household surface had significantly higher levels of H5 HA IgA and IgG (*p*<0.01, respectively).

To confirm these crude findings, we used generalized linear models (GLM) with a generalized estimating equation (GEE) to adjust for clustering at the participant level. Up to 12 models were constructed for each independent variable of interest (PPI, SPI, exposure group, and presence and quantity of Pig-2-Bac DNA) and model fit statistics (QIC, QICu) were compared for covariates and for combinations of covariates including age, sex, household size, season, month of sample collection, having received the current flu vaccine, and occupational livestock exposure. In general, univariate models and models with the least number of covariates resulted in the best model fit (**Tables S1 & S2**). **Table 2** and **Table 3** summarize unadjusted associations and associations adjusted for age and influenza vaccination status. The model estimates confirm the crude associations presented in Figures 1-5 above, e.g., higher poultry and swine production intensity were consistently associated with higher H5 and H1 HA IgA, including after adjusting for participant age and seasonal influenza vaccination status. Higher SPI and PPI were also associated with significantly higher IgG responses to H5 HA for all livestock intensity metrics except for SPI, log_10_. Adjusting for livestock contact instead of excluding those with livestock contact (and thus maintaining the sample size) also showed significantly higher H5 IgG levels with higher PPI and SPI (**Table S4**). Analogously, presence and quantity of Pig-2-Bac (**Table S5**) on home surfaces was associated with significantly higher H5 HA IgA and IgG responses, both in the univariate models and in models adjusted for age and vaccination status (*p*<0.05; **Table 2**).

**Table 2.**
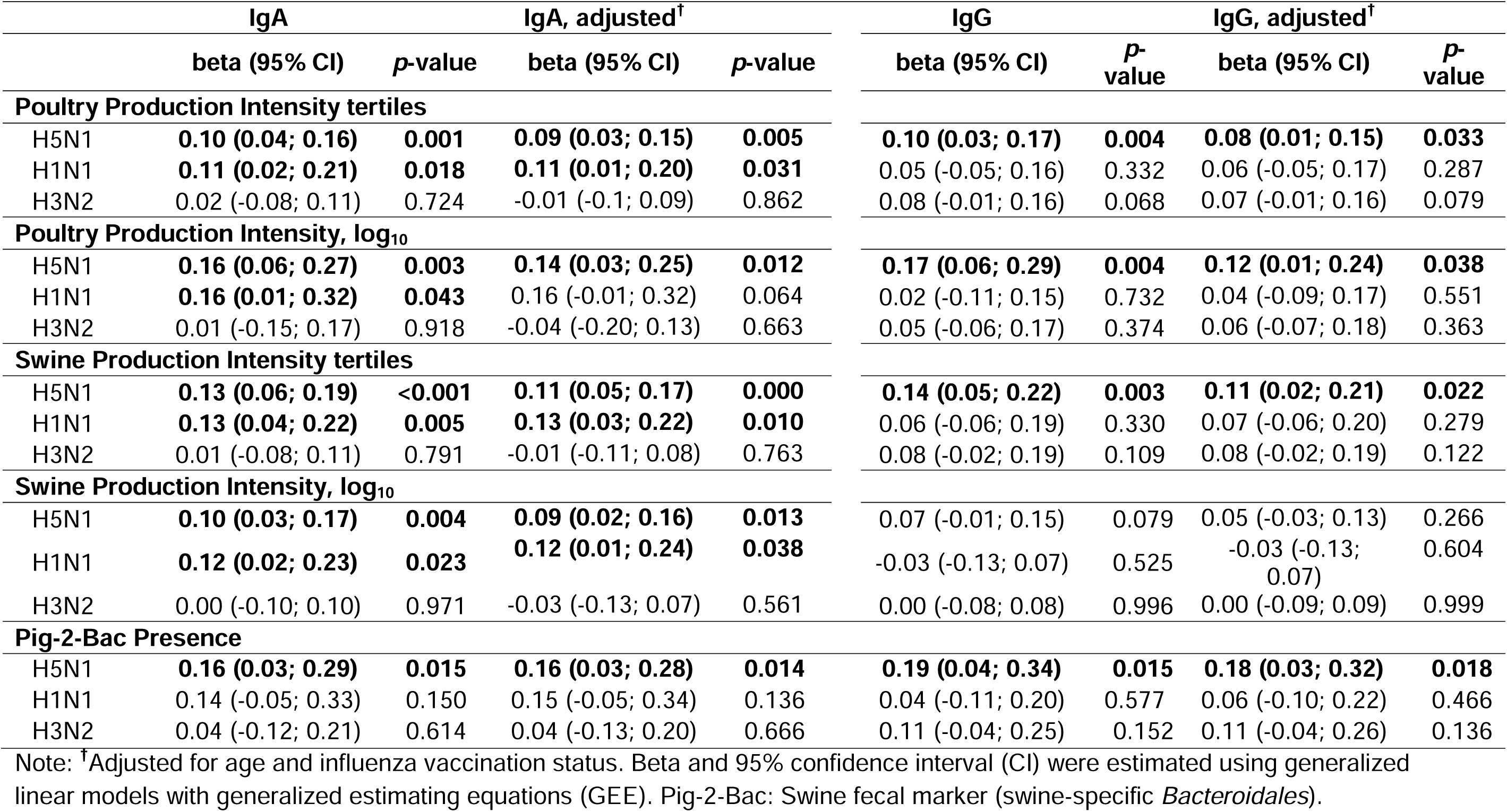
Unadjusted and adjusted (age and influenza vaccination status) associations between poultry and swine production intensity and influenza A antibody responses.

**Table 3.** Unadjusted and adjusted (age and influenza vaccination status) associations between exposure group and influenza A antibody responses.

|  | IgA |  | IgA, adjusted<sup>†</sup> |  | IgG |  | IgG, adjusted<sup>†</sup> |  |
| --- | --- | --- | --- | --- | --- | --- | --- | --- |
|  | beta (95% CI) | p-value | beta (95% CI) | p-value | beta (95% CI) | p-value | beta (95% CI) | p-value |
| **H5N1** |  |  |  |  |  |  |  |  |
| Metro | (Reference) |  | (Reference) |  | (Reference) |  | (Reference) |  |
| ILO-N | **0.16 (0.03; 0.28)** | **0.012** | 0.12 (-0.01; 0.25) | 0.073 | 0.00 (-0.15; 0.15) | 0.998 | -0.07 (-0.21; 0.06) | 0.291 |
| Hog | **0.19 (0.03; 0.35)** | **0.018** | **0.18 (0.01; 0.34)** | **0.035** | **0.21 (0.04; 0.39)** | **0.016** | **0.18 (0.01; 0.34)** | **0.038** |
| Poultry | **0.24 (0.09; 0.40)** | **0.002** | **0.21 (0.05; 0.37)** | **0.009** | **0.34 (0.19; 0.48)** | **<0.001** | **0.35 (0.20; 0.50)** | **<0.001** |
| **H1N1** |  |  |  |  |  |  |  |  |
| Metro | (Reference) |  | (Reference) |  | (Reference) |  | (Reference) |  |
| ILO-N | **0.20 (0.03; 0.36)** | **0.021** | 0.17 (-0.02; 0.37) | 0.082 | -0.04 (-0.21; 0.13) | 0.624 | -0.07 (-0.27; 0.13) | 0.468 |
| Hog | **0.23 (0.03; 0.44)** | **0.026** | 0.21 (-0.01; 0.42) | 0.058 | 0.03 (-0.16; 0.23) | 0.757 | 0.18 (-0.06; 0.41) | 0.136 |
| Poultry | **0.46 (0.26; 0.65)** | **<0.001** | **0.44 (0.24; 0.64)** | **<0.001** | 0.10 (-0.10; 0.29) | 0.335 | -0.12 (-0.41; 0.18) | 0.448 |
| **H3N2** |  |  |  |  |  |  |  |  |
| Metro | (Reference) |  | (Reference) |  | (Reference) |  | (Reference) |  |
| ILO-N | -0.01 (-0.18; 0.16) | 0.938 | -0.09 (-0.28; 0.10) | 0.348 | -0.01 (-0.16; 0.14) | 0.891 | 0.06 (-0.11; 0.22) | 0.513 |
| Hog | -0.04 (-0.24; 0.15) | 0.656 | -0.07 (-0.26; 0.12) | 0.487 | 0.06 (-0.13; 0.24) | 0.550 | **0.24 (0.00; 0.48)** | **0.046** |
| Poultry | **0.31 (0.09; 0.53)** | **0.006** | **0.27 (0.06; 0.48)** | **0.011** | **0.20 (0.04; 0.37)** | **0.013** | 0.06 (-0.18; 0.31) | 0.616 |

Note: <sup>†</sup>Adjusted for age and influenza vaccination status. Beta and 95% confidence interval (CI) were estimated using generalized linear models with generalized estimating equations (GEE).

Comparisons between the ILO-N and ILO-W groups with the Metro group (reference group; **Table 3**) also confirmed crude trends shown in **Figure 4**. The ILO-N group had significantly elevated H5 and H1 HA IgA levels compared to the Metro group, although the association decreased somewhat after adjusting for age and vaccination status. Poultry worker households had the highest H5, H1, and H3 IgA and H5 IgG levels, but hog/pork processing worker households also had significantly elevated H5 HA IgA and IgG and H1 HA responses, which decreased slightly after adjusting for age and vaccination status. Trends shown here were also confirmed in additional models adjusting for age and sampling month (continuous), for age and sampling season, for age, sex and household size (each with GEE component) and were found to remain robust. Additionally, outcomes were confirmed with GEE clustering at the household level instead of clustering at the participant levels, which did not also affect the main trends described above.

**Figure 4.**
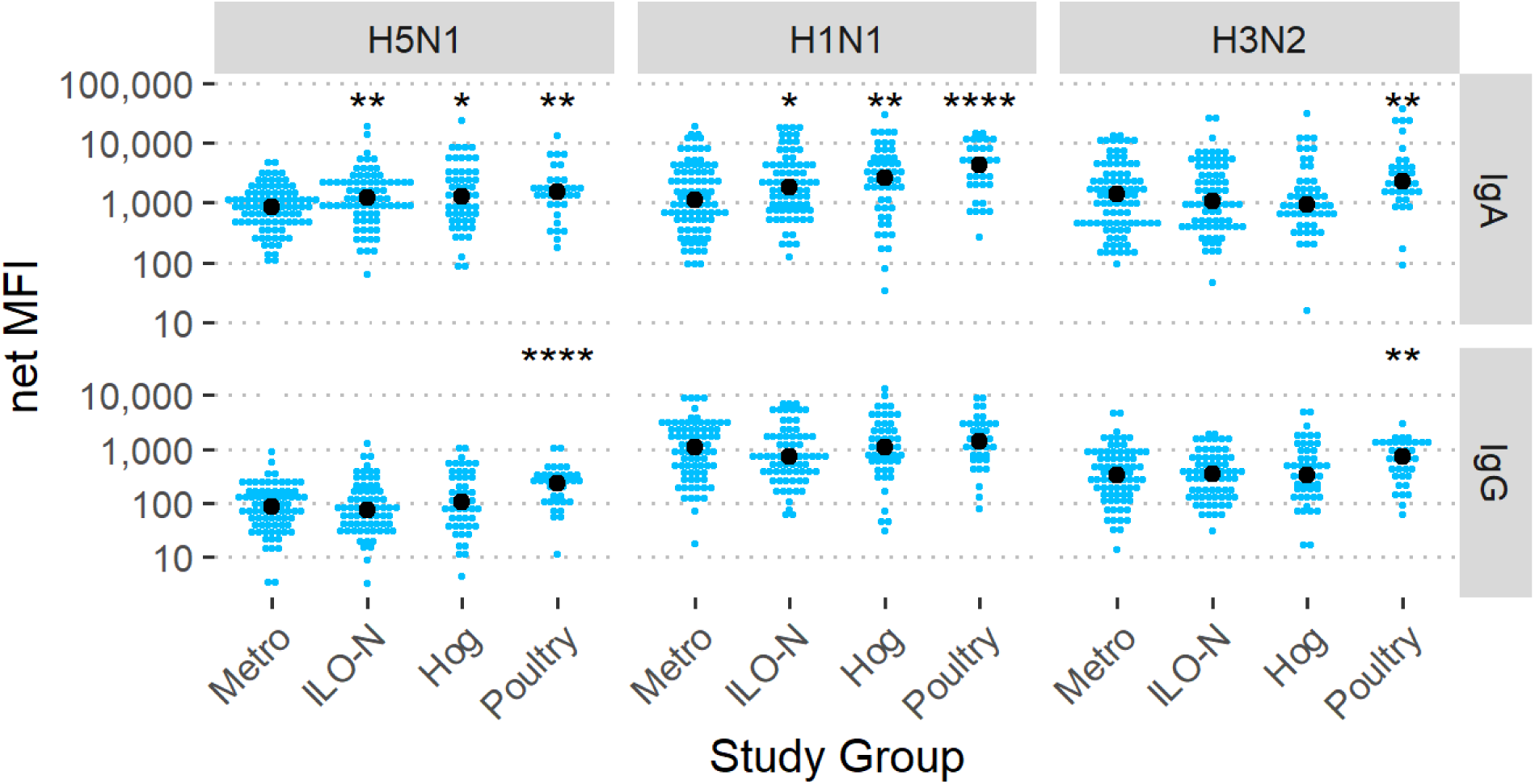
IgA and IgG antibody levels specific to influenza A H1 HA, H3 HA, and H5 HA stratified by livestock exposure group. Note: *=p<0.05, ** = p<0.01, *** = p<0.001, ****=p<0.0001. Reference group for statistical comparison: Metro. ILO-N: Industrial Livestock Operation Neighbor Households, Hog/Poultry: Households with 1 or more household members working at Industrial Hog (Hog) or Poultry Operations (Poultry). MFI: Median Fluorescence Intensity

**Figure 5.**
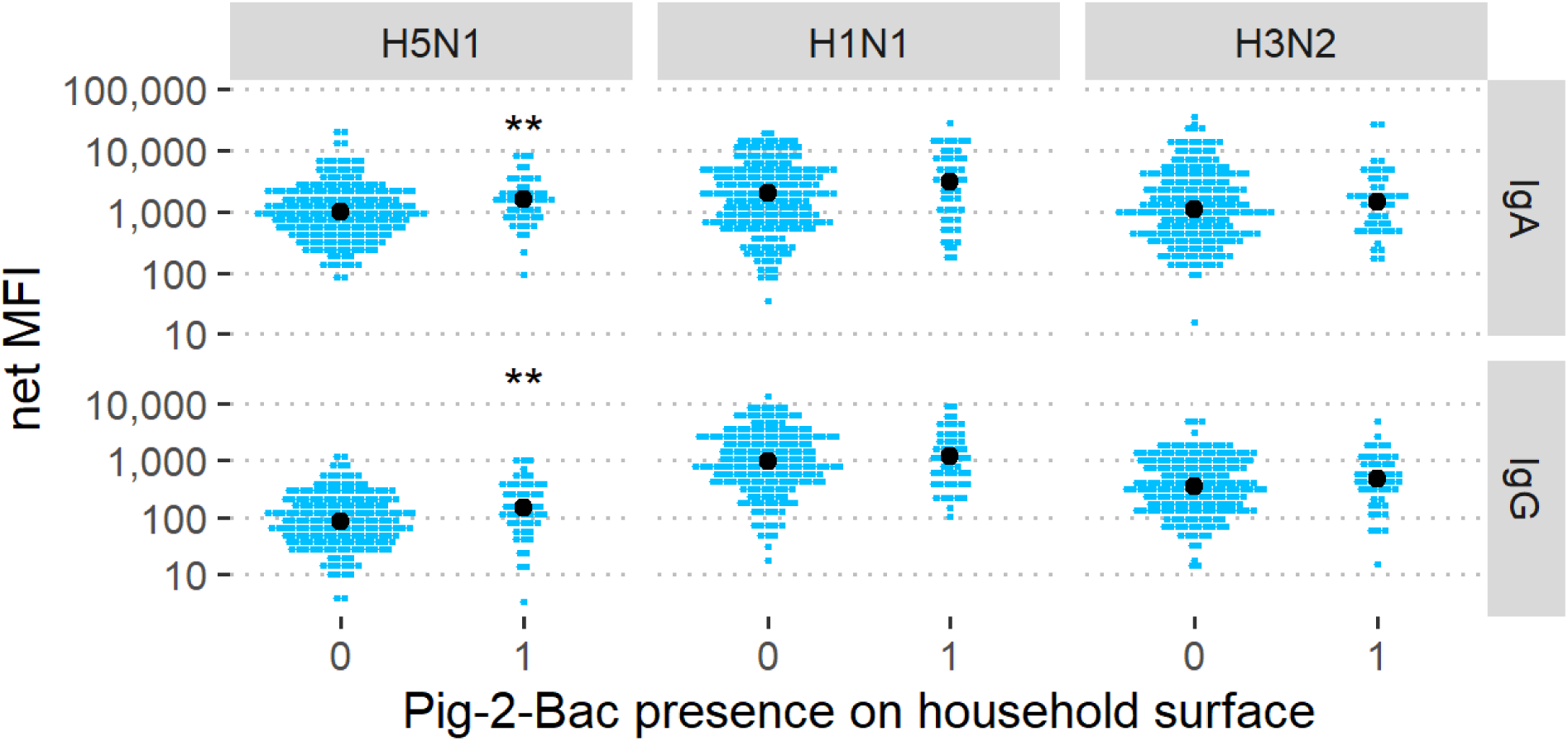
Influenza A antibody levels stratified by presence of Pig-2-Bac, a swine-specific microbial source tracking marker. Note: *=p<0.05, ** = p<0.01, *** = p<0.001. Reference group for comparison: 0 (no Pig-2-Bac DNA detected). MFI: Median Fluorescence Intensity

## Discussion

Our analysis of saliva samples from a study conducted during the COVID-19 pandemic (February 2021 to December 2022) in North Carolina, showed significantly elevated levels of antibodies to H5 HA among community members living in areas of high poultry and swine production intensity, including among residents who were not occupationally exposed to livestock and who did not have any household members who worked with livestock or at meat processing plants. ILO workers and their household members without direct occupational livestock exposure had the highest levels of H5 HA IgA and IgG and H1 HA IgA compared to ILO-N and to Metro households, but ILO-N households without direct livestock exposure also had significantly elevated H5 HA and H1 HA IgA levels compared to the Metro group. ILO worker households that had at least one family member exposed to live or dead poultry through work at industrial poultry operations or poultry meat processing plants had the highest levels of H5, H1, and H3 HA IgA, and of H5 and H3 HA IgG. North Carolina experienced 14 poultry HPAI outbreaks in 2022, including in counties from where this study enrolled participants. Most impacted were the commercial broiler production and commercial turkey meat birds with more than 300,000 broilers and nearly 75,000 turkeys affected by the end of 2022 [21]. Most ILO workers in this study were employed in animal slaughtering and meat processing plants [16]. The high H5, H1 and H3 IgA levels among poultry workers suggest that poultry workers could have been involved in the disposal of infected birds or in the inspection of infected birds.

Our prior research in this community showed that swine fecal contamination of household surfaces (presence and quantity of Pig-2-Bac DNA) increased significantly with local swine production intensity [17] and discussed how swine fecal bacteria can indicate the presence of other potentially pathogenic zoonotic microorganisms, including those with pandemic potential. Here we add another layer of research beyond the environmental contamination of home surfaces with swine fecal material by demonstrating that the presence and quantity of swine fecal contamination on community homes (mean copy numbers of Pig-2-Bac DNA per m^2^ of home surface tested) were associated with significantly higher levels of mucosal H5 HA IgA. Although pigs are not very susceptible to severe H5N1 [22,23], they could be carriers or act as mixing vessels since they have receptors for both the human and avian influenza viruses. In regions like eastern NC with high co-location of swine and poultry production, this could facilitate genetic reassortment and the emergence of novel pandemic influenza strains. Since swine typically do not show symptoms of H5N1 infection, pigs carrying H5N1 asymptomatically may not be identified and enter the slaughtering and meat processing pipeline undetected [24].

These findings contrast with recent studies that found that baseline immunity to contemporary clade 2.3.4.4b H5N1 influenza viruses in humans is limited and with statements by federal institutions that maintain that the threat of contracting H5N1 to the general public remains low [25]. Two recent serological studies reported low to minimal levels of cross-reactive antibodies to bovine H5N1 hemagglutinin (HA) antigens of clade 2.3.4.4b in two urban populations between 2024 and 2025 [26,27]. Both studies showed that population immunity to clade 2.3.4.4b H5N1 viruses is dominated by neuraminidase (NA)-directed antibody responses (i.e., against N1), whereas HA-directed responses (i.e. against H5) remain low or absent [27]. While seasonal vaccination did not boost H5N1 antibody levels, influenza A H1N1 infection boosted HPAI N1 NA antibodies [26]. The lack of H5 HA antibodies in urban populations is consistent with our findings, i.e., we observed the lowest H5 HA IgG (and IgA) binding antibody levels in our Metro group, but significantly and consistently higher H5 HA antibody levels across a variety of livestock exposure metrics, including poultry and swine production intensity around the residence, living in a household with an individual with occupational exposure to poultry or swine, and detection and quantity of swine fecal material on household surfaces.

While it is possible that this study measured cross-reactive H5 HA IgA and IgG, e.g., against H5NX (where NX = N2, N3, etc.) rather than HPAI H5N1 HA binding antibodies, our findings nonetheless indicate a zoonotic pathogen source demonstrated by positive associations between H5 HA antibody levels with all parameters explored (occupational exposure, residential proximity, livestock fecal microbial source tracking marker). Other limitations include the absence of pathogen detection directly or documentation of active H5N1 infection, e.g., via influenza A virus RT-qPCR from repeated air or household surface sampling or from repeated participant nasal swab sampling during the study period. While feasible, this approach would likely require a larger cohort with longitudinal sampling in each group, as influenza virus infections are typically only detected during a short acute phase window and, in a symptom-triggered study design, could be missed completely if the infection is mild or asymptomatic. Serological approaches represent an integrated solution that allows for detection of both recent (IgA) and past influenza infection (IgG), thereby simplifying study protocols, study costs and burden on the participants – important considerations for work like this where community members contribute to the design, conduct, and dissemination of the study.

Influenza viruses can spread from infected animals into the environment (and onto nearby household surfaces) through wind or via aerosol dispersion [28,29]. Additionally, influenza viruses can spread through feathers generated during the culling of infected birds, decomposing buried carcasses, and flies transmitting the virus to their next target after feasting on infected or rotting birds or bird feces. Duck farm density and proximity to poultry farms were other risk factor for HPAI spread identified in France [30], similar to our findings among communities living near such farms. France has since started to vaccinate its ducks against H5N1. Influenza A viruses can also spread through runoff or aerosols from swine manure applied to crop fields or from poultry litter applied to fields as fertilizer, as suggested by the detection of bacteria from pig fecal waste (Pig-2-Bac DNA) on ILO-N homes. Viruses are much smaller than bacteria and likely can spread much farther than potentially pathogenic bacteria from animal wastes; up to ∼5 miles (8 km) according to a Czech investigation [28]. Another potential exposure route in ILO worker households is by taking home clothes, masks, shoes, or personal protective equipment (PPE) worn during work at livestock operations or in meat processing plants where infected animals or animal parts were present, inspected, processed, or culled and buried or otherwise disposed of. Take-home exposure from dairy workers in contact with positive dairy herds – perhaps via contaminated clothing (fomite-based transmission) – is suspected in the cases of several indoor cats that died from HPAI H5N1 infection [31].

Data from this study suggest that H5N1 exposure with either mild symptoms that do not warrant emergency visits or hospitalization or asymptomatic infection could be common among community members living in areas of high livestock production intensity, and that livestock worker households, particularly poultry worker households, bear the brunt of these exposures. These findings also suggest that industrial livestock operations, including concentrated animal feeding operations (CAFOs) and meat processing plants, represent significant sources of zoonotic infectious disease spillover and risk of infection for ILO workers and for community members living nearby, in particular for residents living downwind from these operations or from crop and sprayfields.

Future research should include development and employment of poultry-specific microbial source tracking markers to investigate whether homes located in areas of high poultry production intensity are contaminated with chicken and turkey fecal material and other zoonotic microbes, including H5N1 virus that may be present in poultry fecal material. There are several established poultry markers, including Av4143, which detects a bird fecal *Lactobacillus* present in multiple avian species, and cytB, a chicken and duck mitochondrial (cellular) marker [32,33]. However, neither of these markers is specific to poultry (chicken and turkey) fecal material exclusively, which are of most concern in this region. While additional research is needed to determine whether H5N1 exposure and infection risk among communities living in areas of intensive and often co-located poultry and swine production have changed since this study was conducted, these findings warrant increased surveillance of those living and working at the interface of industrial livestock production and meat processing. Ideally, the option of anonymous serological and antigen testing — e.g., through community engagement — could be provided in these settings to help inform communities of risks, offer self-testing kits, and to protect ILO workers from potential loss of employment or reprimand for seeking clarity around H5N1 and other zoonotic infection risks due to working with livestock. Lastly, these findings should be considered in ongoing discussions about whether H5N1 vaccination should be offered to workers at high risk of infection and whether poultry vaccination could help limit spread between farms and between animals and people living in areas of intensive livestock production.

## Data Availability

All data produced in the present study are available upon reasonable request to the authors.

## Acknowledgements

We would like to thank the community organizers at the Rural Empowerment Association for Community Help (REACH), especially Margaret Carr, Clesha Hall, Unique Hall, Angela Matthews, Arika Miller, Helen Santizo, Phyla Holmes, Norma Mejia, and Sherneka Smith. This study would not have been possible without the strong partnership that REACH has with members of communities in areas of dense industrial hog production. We would like to thank all the participants who agreed to be part of this study.

