## Supplement for "Livestock production intensity and mucosal IgA and IgG responses to H5N1 highly pathogenic avian influenza A virus, North Carolina, 2021-2022"

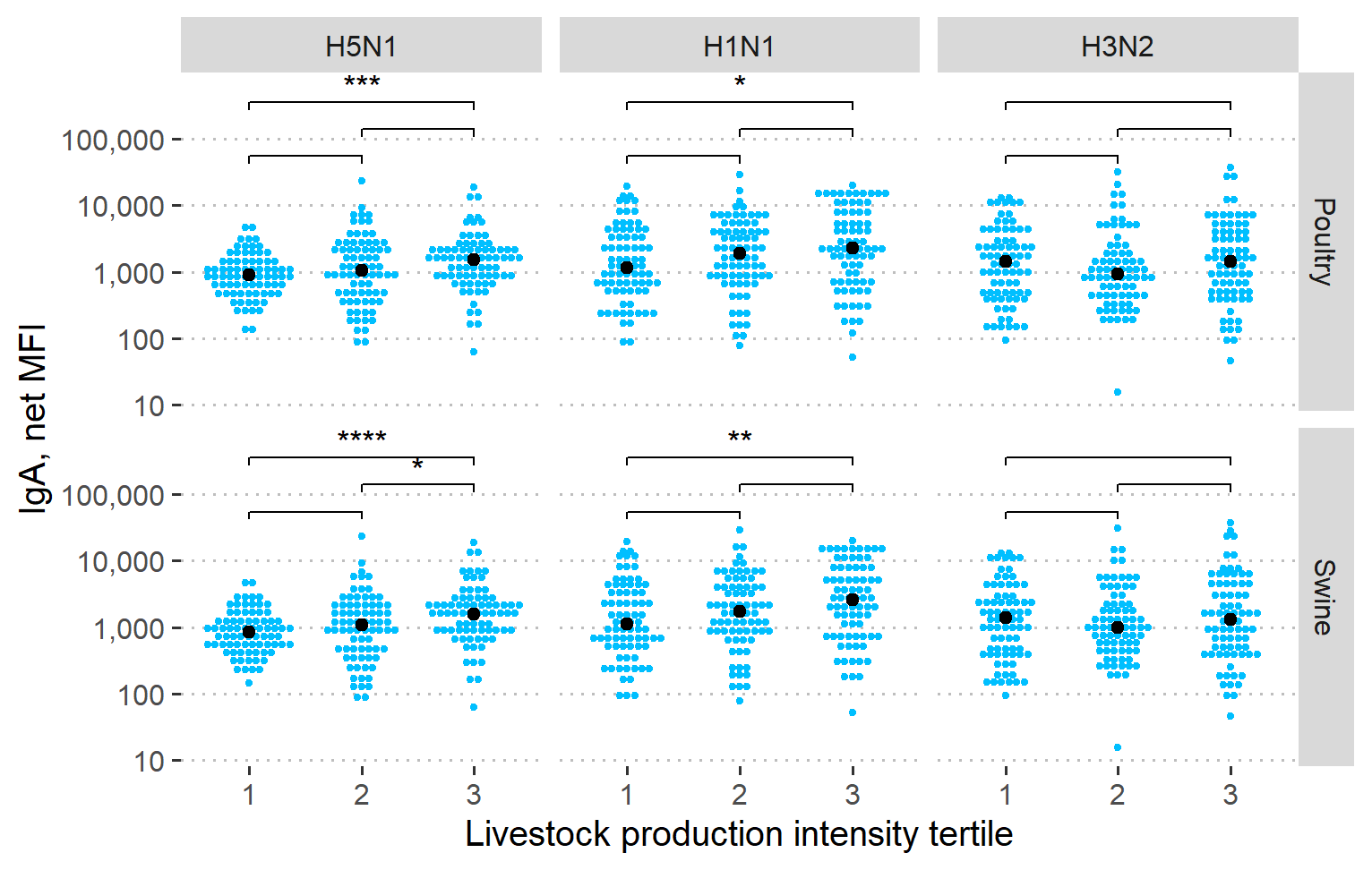

Figure S1. Influenza A IgA response including children (<18 years, n = 13) by livestock production intensity tertile. Note: *=p<0.05, ** = p<0.01, *** = p<0.001, ****=p<0.0001. Reference group for statistical tests is tertile 1. MFI: Median Fluorescence Intensity.

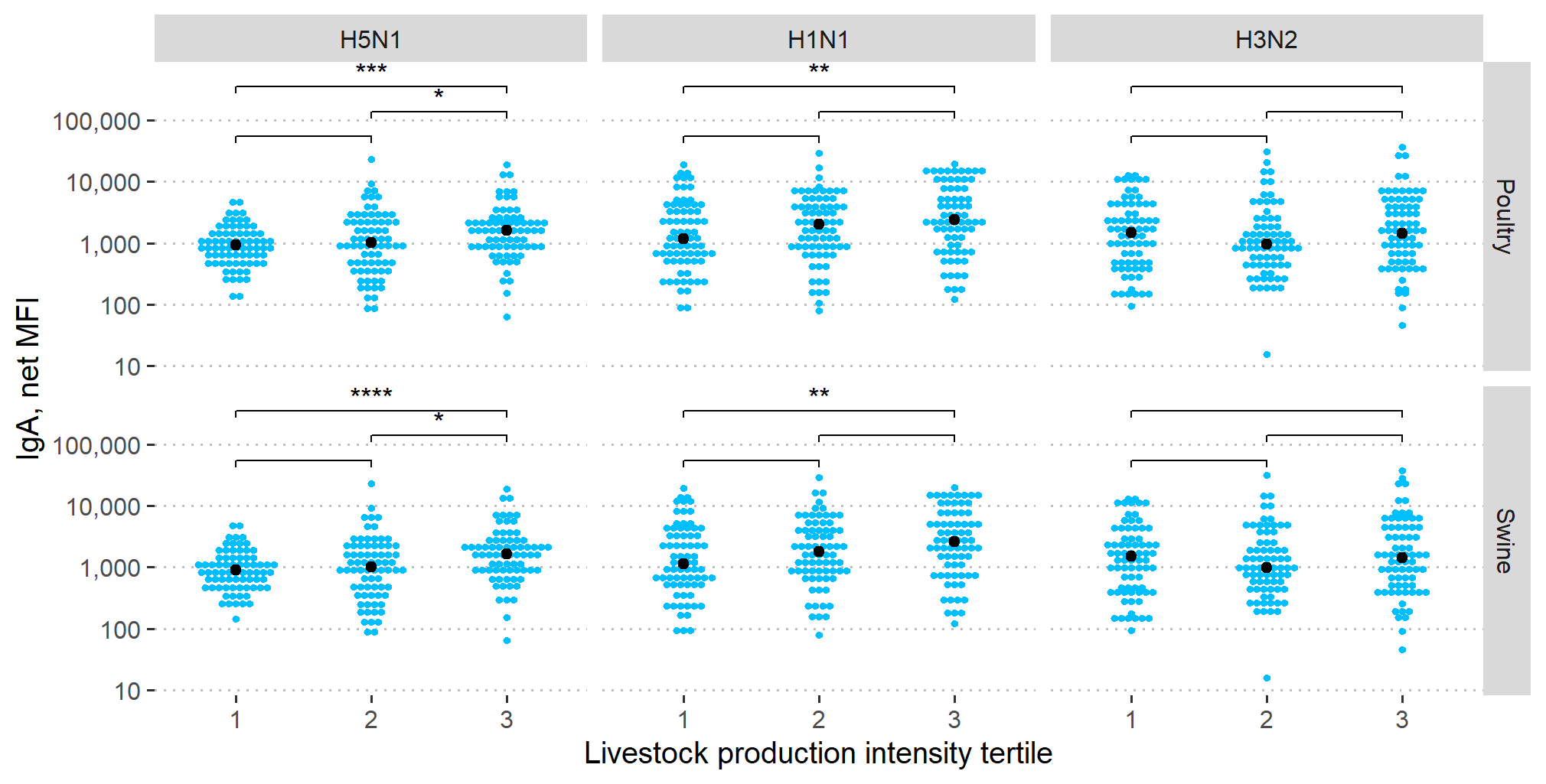

Figure S2. IgA responses to influenza A H1N1, H3N2, and H5N1 by livestock production intensity tertile. Children (<18 years) were excluded from this analysis. Note: *=p<0.05, ** = p<0.01, *** = p<0.001, ****=p<0.0001. MFI: Median Fluorescence Intensity.

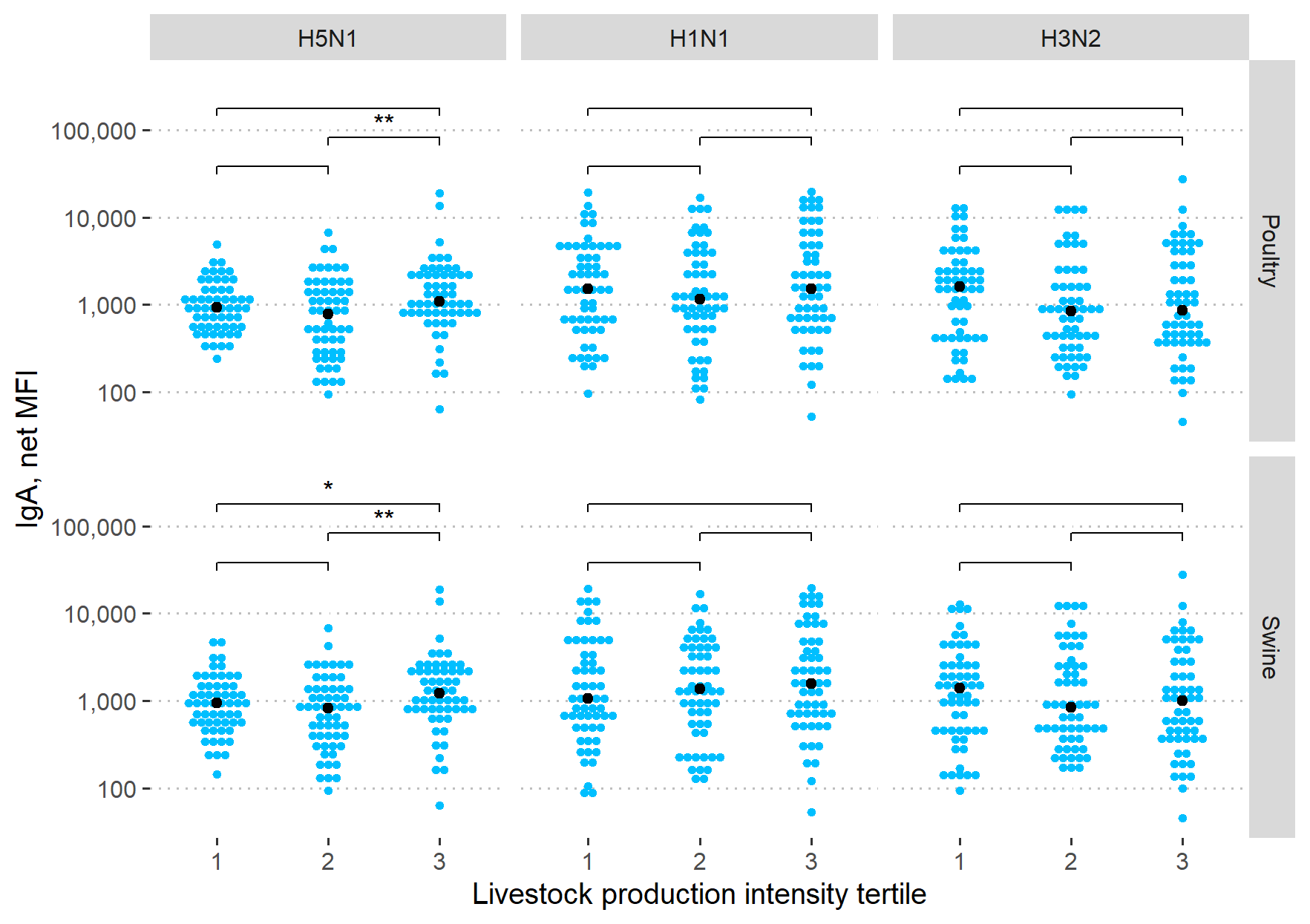

Figure S3. IgA to H5N1 among community members without occupational livestock contact. Note: *=p<0.05, ** = p<0.01, *** = p<0.001, ****=p<0.0001. MFI: Median Fluorescence Intensity.

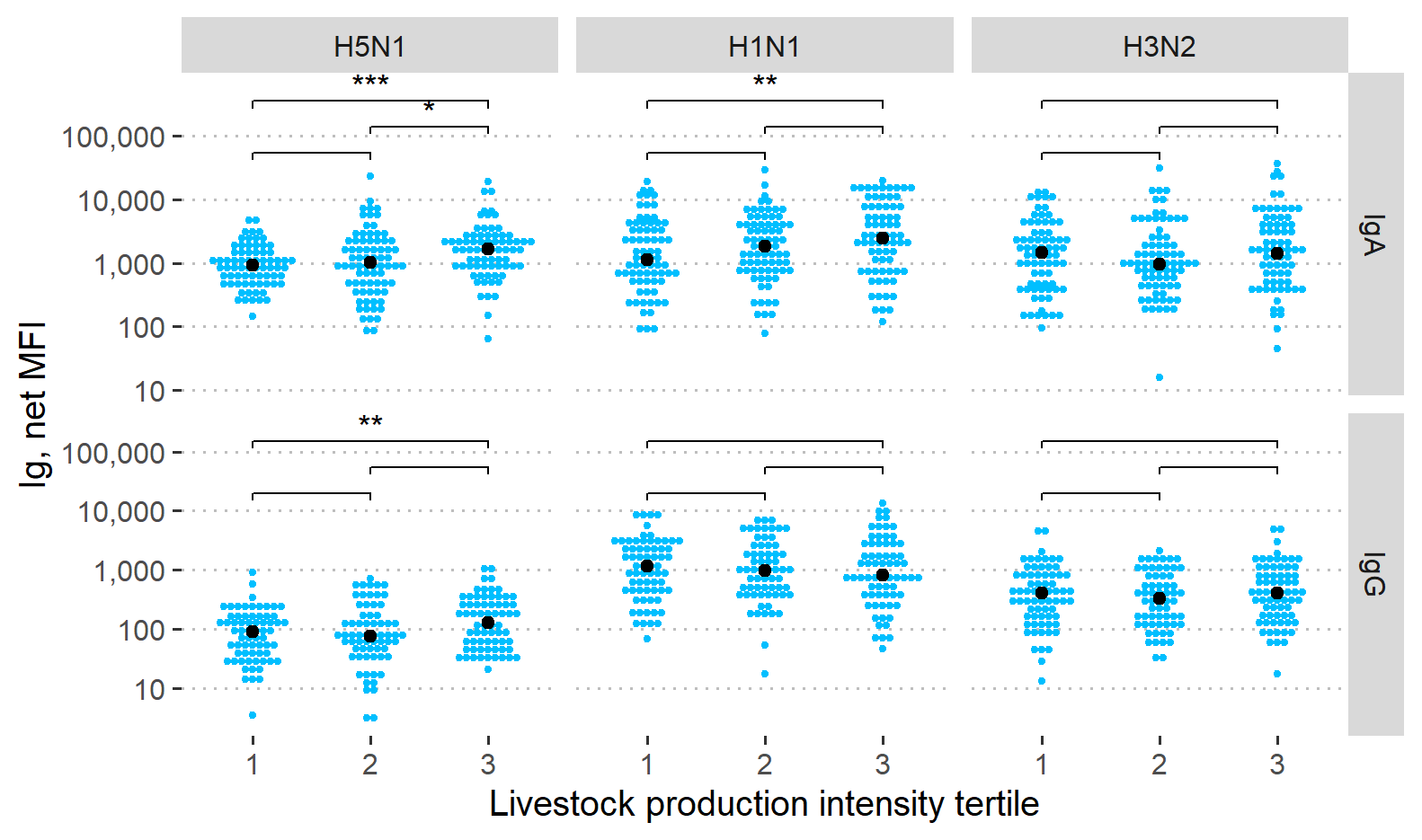

Figure S4. Influenza A antibody responses by swine and poultry production intensity tertile combined. Note: *=p<0.05, ** = p<0.01, *** = p<0.001, ****=p<0.0001. MFI: Median Fluorescence Intensity.

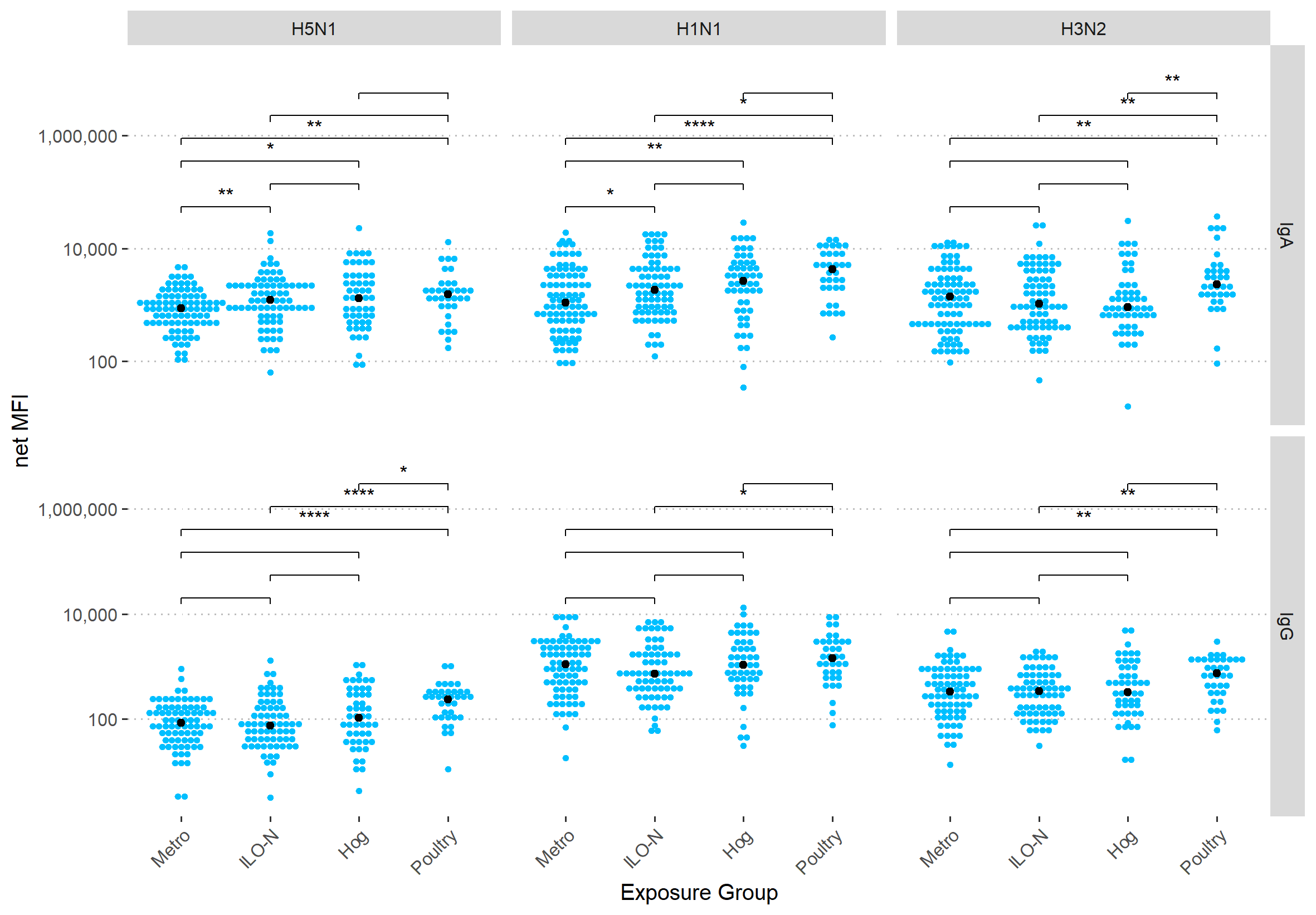

Figure S5. IgA and IgG antibody levels specific to influenza A H1N1, N3N2, and H5N1 stratified by livestock exposure group. Note: *=p<0.05, ** = p<0.01, *** = p<0.001, ****=p<0.0001. Associations remain unchanged when children (<18 years, n = 13) are included in the analysis. MFI: Median Fluorescence Intensity.

**Table S 1.** Comparison of model fit statistics for unadjusted versus different covariate adjustment sets. Independent variable: Poultry Production Intensity (log_10_); Dependent variable: H5N1 IgA (log_10_).

| **Covariates** | **QICu** | **Delta** | **Weight** |
| --- | --- | --- | --- |
| Univariate (none) | 217.99 | 0.00 | 0.305 |
| Livestock contact (binary) | 219.97 | 1.99 | 0.113 |
| Time (months, continuous) | 219.98 | 1.99 | 0.113 |
| Age | 219.98 | 1.99 | 0.113 |
| Sex | 219.98 | 1.99 | 0.113 |
| Household size | 219.99 | 2.00 | 0.112 |
| Age + sex | 221.98 | 3.99 | 0.041 |
| Flu vaccine | 221.99 | 4.00 | 0.041 |
| Age + sex + household size | 223.98 | 5.99 | 0.015 |
| Season | 223.98 | 5.99 | 0.015 |
| Age + flu vaccine | 223.98 | 5.99 | 0.015 |
| Age + sex + household size + flu vaccine | 227.98 | 9.99 | 0.002 |

**Table S 2.** Comparison of model fit statistics for unadjusted versus different covariate adjustment sets. Independent variable: Mean Pig-2-Bac copy number per m^2^(log_10_); Dependent variable: H5N1 IgA (log_10_).

| **Covariates** | **QICu** | **Delta** | **Weight** |
| --- | --- | --- | --- |
| Univariate (none) | 230.99 | 0.00 | 0.30 |
| Age | 232.93 | 1.93 | 0.11 |
| Livestock contact (binary) | 232.94 | 1.95 | 0.11 |
| Time (months, continuous) | 232.97 | 1.97 | 0.11 |
| Sex | 232.99 | 1.99 | 0.11 |
| Household size | 232.99 | 2.00 | 0.11 |
| Flu vaccine | 234.92 | 3.93 | 0.04 |
| Age + sex | 234.93 | 3.94 | 0.04 |
| Age + flu vaccine | 236.85 | 5.86 | 0.02 |
| Age + sex + household size | 236.93 | 5.94 | 0.02 |
| Season | 236.98 | 5.99 | 0.02 |
| Age + sex + household size + flu vaccine | 240.85 | 9.86 | 0.00 |

**Table S 3.** Association between poultry and swine production intensity (PPI, SPI tertiles) and influenza A antibody response among community members with no occupational exposure to livestock, North Carolina, USA, 2021-2022.

|  | **IgA** |  | **IgG** |  |
| --- | --- | --- | --- | --- |
|  | **beta (95% CI)** | ***p*-value** | **beta (95% CI)** | ***p*-value** |
| **PPI tertiles** | |  |  |  |
| H5N1 | **0.10 (0.03; 0.16)** | **0.006** | 0.04 (-0.04; 0.12) | 0.334 |
| H1N1 | 0.06 (-0.04; 0.17) | 0.211 | -0.03 (-0.13; 0.07) | 0.520 |
| H3N2 | -0.01 (-0.12; 0.09) | 0.841 | 0.00 (-0.08; 0.07) | 0.904 |
| **SPI tertiles** | | |  |  |
| H5N1 | **0.11 (0.04; 0.17)** | **0.003** | 0.05 (-0.03; 0.13) | 0.235 |
| H1N1 | 0.08 (-0.02; 0.18) | 0.112 | -0.03 (-0.13; 0.07) | 0.586 |
| H3N2 | -0.02 (-0.13; 0.09) | 0.696 | -0.01 (-0.09; 0.07) | 0.831 |

*Note. Beta and 95% confidence interval (CI) were estimated using generalized linear models with generalized estimating equations (GEE). SE: standard error.*

**Table S 4.** Association between poultry and swine production intensity (PPI, SPI tertiles) and influenza A antibody responses adjusting for livestock, North Carolina, USA, 2021-2022.

|  | **IgA** |  | **IgG** |  |
| --- | --- | --- | --- | --- |
|  | **beta (95% CI)** | ***p*-value** | **beta (95% CI)** | ***p*-value** |
| **PPI tertiles adjusted for livestock contact** | | | | |
| H5N1 | **0.08 (0.01; 0.14)** | **0.026** | **0.09 (0.02; 0.15)** | **0.013** |
| H1N1 | 0.08 (-0.07; 0.23) | 0.287 | 0.04 (-0.07; 0.14) | 0.487 |
| H3N2 | -0.01 (-0.12; 0.09) | 0.780 | 0.06 (-0.01; 0.14) | 0.111 |
| **SPI tertile adjusted for livestock contact** | | | | |
| H5N1 | **0.10 (0.04; 0.17)** | **0.003** | **0.10 (0.02; 0.18)** | **0.011** |
| H1N1 | 0.1 (-0.05; 0.25) | 0.197 | 0.04 (-0.08; 0.16) | 0.501 |
| H3N2 | -0.01 (-0.12; 0.09) | 0.836 | 0.06 (-0.03; 0.16) | 0.174 |

**Table S 5.** Association between Pig-2-Bac quantity on residents’ household surfaces (Pig-2-Bac copy number/m^2^) and influenza A antibody response, North Carolina, USA, 2021-2022.

|  | **IgA** |  | **IgG** |  |
| --- | --- | --- | --- | --- |
|  | **beta (95% CI)** | ***p*-value** | **beta (95% CI)** | ***p*-value** |
| **Pig-2-Bac/m^2^** |  |  |  |  |
| H5N1 | **0.06 (0.01; 0.12)** | **0.022** | **0.07 (0.02; 0.13)** | **0.014** |
| H1N1 | 0.04 (-0.04; 0.13) | 0.300 | 0.03 (-0.04; 0.09) | 0.434 |
| H3N2 | 0.02 (-0.05; 0.09) | 0.604 | 0.05 (-0.02; 0.11) | 0.144 |

Note. Beta and 95% confidence interval (CI) were estimated using generalized linear models with generalized estimating equations (GEE). SE: standard error.
